# Stage Slip from Diagnostic Latency in MCED Trials: A Calibrated Monte Carlo Reconstruction of the NHS-Galleri Results

**DOI:** 10.64898/2026.03.01.26347360

**Authors:** Hamid Bellout

## Abstract

**Background:** The NHS-Galleri trial reported a substantial reduction in Stage IV cancer diagnoses and a four-fold increase in cancer detection rates, but did not meet its primary endpoint of reducing combined Stage III+IV diagnoses in a prespecified group of 12 cancers. We hypothesize that *stage slip*— progression of cancers from Stage I/II to Stage III during diagnostic workup—is the primary mechanism behind this statistical masking.

**Methods:** We developed a Monte Carlo simulation of 142,000 participants (matching NHS-Galleri enrolment) across 12 cancer types, calibrated to NHS England population stage distributions. The model represents three competing clocks: biological sojourn times, screening-initiated or symptom-driven pathway initiation, and diagnostic infrastructure delays. The median standard-of-care diagnostic delay (92 days) was constructed from five convergent evidence streams. We estimated the number of intervention-arm cases where a cancer was biologically Stage I/II but recorded as Stage III+IV at diagnosis, and determined how many such cases would need to be *recovered* for the composite endpoint to reach statistical significance. We validated that the 12 deadly cancers produce a sufficiently large early-stage patient pool and confirmed robustness across a wide range of test sensitivity and diagnostic delay assumptions.

**Findings:** The calibrated control arm reproduces NHS stage distributions within 1–2 percentage points for all 12 cancers. In the intervention arm, we estimate 84 cases of stage slip (95% CI: 68–104) at published sensitivity values. If only 25 of these cases (approximately one in three) had been diagnosed before crossing the Stage II/III boundary, the composite endpoint would have reached *p <* 0.05 at reported sensitivity levels. The slip count is robust to sensitivity assumptions, varying from approximately 74 to 85 cases across a 30–100% range of published CCGA 3 values. Across diagnostic delays from 65 to 120 days (at realised sensitivity), the estimate ranges from approximately 55 to 95 cases. The expected pool of early-stage cancers in the 12 deadly types (∼ 260–335 screen-detected Stage I/II cases per arm) is large enough that even a 12% slip rate—corresponding to the most optimistic delay assumption—produces sufficient slippage to exceed the significance threshold.

**Interpretation:** Stage slip provides a quantitatively sufficient and mechanistically transparent explanation for the primary endpoint miss. The test’s biological performance is intact: it detects cancers earlier and reduces Stage IV diagnoses. The composite endpoint was attenuated by systemic diagnostic latency in the NHS, not by a failure of the assay. Future MCED trials should consider endpoints less vulnerable to infrastructure delays or incorporate prespecified adjustments for expected diagnostic pathway timing.

**Funding:** None.

**Registration:** Not applicable.

## 1 Introduction

Multi-cancer early detection (MCED) tests aim to shift the stage distribution of cancers toward earlier, more treatable stages. The NHS-Galleri trial, the largest randomized controlled trial of an MCED test to date, enrolled 142,000 participants in England’s National Health Service and evaluated annual screening with the Galleri blood test over three years [1]. The trial’s primary objective was to demonstrate a statistically significant reduction in late-stage (Stage III+IV) cancer diagnoses, assessed sequentially across three prespecified cancer groupings. The first gate in this sequential analysis — and the one on which the trial’s significance depended — was a prespecified group of 12 cancer types that account for approximately two-thirds of cancer deaths in the UK and US, most of which lack any existing screening programme

Public reporting indicated substantial positive signals: a reduction in Stage IV diagnoses exceeding 20% in screening rounds 2 and 3, a four-fold improvement in overall cancer detection rates, and a substantial increase in Stage I/II detections [1]. However, the primary composite Stage III+IV endpoint was not met (consistent with a p-value in the range 0.05–0.20), with the sponsor noting “a higher than anticipated incidence of Stage III cancers” [1].

This study addresses a specific question: is the endpoint miss explained by diagnostic infrastructure timing rather than test failure? A composite Stage III+IV endpoint counts cancers by stage *at diagnosis*, not by stage *at pathway initiation*. If a screening signal triggers diagnostic workup while a cancer is Stage I/II but confirmation occurs after the cancer has crossed the Stage II/III boundary, the case is recorded as Stage III—*against* the endpoint the test is trying to reduce. We term this mechanism **stage slip**.

We present a calibrated Monte Carlo simulation framework that quantifies stage slip under explicitly stated assumptions, demonstrates its robustness to key parameter choices, and shows that a modest number of slipped cases is sufficient to explain the observed endpoint miss.

## 2 Methods

### 2.1 Conceptual framework: three competing clocks

Stage slip arises from a race among three temporal processes:

a. **Biological clock (sojourn time)** Each cancer dwells in each stage for a finite, stochastic duration. We model sojourn times as cancer-specific lognormal random variables with literature-informed medians and a shared dispersion parameter *σ* = 0.35.
b. **Signal clock (pathway initiation)** In the intervention arm, the signal is a positive MCED test at an annual screening round. In the control arm, it is the onset of symptoms sufficient to trigger clinical referral, modeled via calibrated stage-dependent exponential hazards. The key asymmetry: Galleri can fire while a cancer is Stage I/II and the patient is asymptomatic; the symptom-driven signal almost never fires at Stage I/II because early-stage cancers are largely asymptomatic.
c. **Infrastructure clock (diagnostic delay)** The elapsed time from signal to confirmed diagnosis. If this exceeds the remaining sojourn time before the Stage II/III boundary, the patient slips.

#### Why slip matters only in the intervention arm

In the control arm, cancers that progress from Stage I/II to III/IV are following their natural history—no early detection event occurred, no accelerated pathway was initiated, and no infrastructure failed to keep pace. This is the baseline rate of late-stage diagnosis in an unscreened population. In the intervention arm, the test detected the cancer early and initiated a diagnostic pathway; if the pathway was too slow, the early detection advantage was lost. Stage slip is therefore an infrastructure cost borne exclusively by the screened arm.

### 2.2 Cohort, cancer mixture, and screening schedule

We simulate a cohort of *N* = 142,000 participants (matching NHS-Galleri enrolment) with annual screening at *t* ∈ {0, 1, 2} years and three-year follow-up. Twelve cancer types are modeled with weights *w*_*c*_ reflecting English population incidence for the relevant age band (Table 10).

#### 2.2.1 Expected case counts

At an approximate annual incidence rate of ∼ 350 per 100,000 for these 12 cancer types in the 50–77 age band, the trial is expected to produce approximately 1,500 cancers across both arms (∼ 750 per arm) over three years. This is a large pool from which stage slip can draw: even a modest slip rate applied to the early-stage fraction produces a clinically consequential number of misclassified cases.

### 2.3 MCED sensitivity: published values for the 12 deadly cancers

A critical input is the Galleri test’s stage-specific sensitivity for each cancer type. The test was clinically validated in the third Circulating Cell-free Genome Atlas (CCGA 3) substudy [2, 3], which reported cancer-specific and stage-specific sensitivities. For the 12 prespecified deadly cancers, the test achieves substantially higher sensitivity than the all-cancer average because these aggressive tumors shed more cell-free DNA at earlier stages [2, 4].

Published data include:

- **Aggregate:** 76.3% sensitivity across all stages for the 12 deadly cancers; 67.6% at Stages I–III [2].
- **Cancer-specific** (from GRAIL HCP documentation [5]): pancreatic cancer 61.9% at Stage I, 60.0% at II, 85.7% at III; ovarian cancer 50.0% at Stage I, 80.0% at II, 87.1% at III; liver/bile duct 100% at Stage I.
- **All-cancer averages** (for comparison): Stage I 16.8%, Stage II 40.4%, Stage III 77.0%, Stage IV 90.1% [2].

The all-cancer averages substantially *understate* sensitivity for the 12 deadly cancers. Our simulation uses cancer-specific values derived from these published sources (Table 10). To verify robustness, we also test a conservative scenario using the lower all-cancer averages (Section 4).

#### 2.3.1 Validation of the early-stage patient pool

With a weighted average Stage I/II sensitivity of approximately 35–45% for these 12 cancers, and ∼ 750 cases per arm, the test is expected to detect 260–335 cancers at Stage I/II in the intervention arm. This pool is large enough that even a 12% slip rate (the most optimistic scenario, corresponding to a 65-day delay) would produce ∼ 36 slipped cases—still above the 25-case significance threshold established in our counterfactual analysis (Section 2.7).

### 2.4 Control-arm calibration to NHS stage distributions

A credible stage-slip estimate requires that the control arm reproduce the actual NHS population stage-at-diagnosis distribution by cancer type. We model symptom-driven referral using stage-dependent exponential hazards *k*_*E,c*_, *k*_III,*c*_, *k*_IV,*c*_ for each cancer. These were calibrated via grid search so that simulated control-arm stage distributions match NHS targets compiled from:

- National Lung Cancer Audit (NLCA) 2022–2023 [11]
- NHS Digital cancer incidence by stage, 2015–2022 [12]
- Cancer Research UK Early Diagnosis Data Hub [10]

Table 1 confirms all 12 cancers match within 1–2 percentage points RMS.

### 2.5 Diagnostic delay model

#### 2.5.1 Construction of the 92-day central estimate

The standard-of-care median diagnostic delay Δ_SoC_ = 92 days is the single most consequential assumption. It is a **constructed estimate** derived from five convergent evidence streams:

i. **PATHFINDER 2 optimised US floor (46 days)**. Median diagnostic resolution in optimised US academic centres; true positives resolved in 36 days (IQR 24–61) [6].
ii. **PATHFINDER 1 early US experience (79 days)**. Before workup protocols were refined; false-positive median was 162 days [7].
iii. **Trial-induced spillover (Mann et al**., **RAND 2025)**. Difference-in-differences analysis across 9.6 million NHS referrals showing a +3.4 percentage point increase in diagnostic delay rates in participating regions (*p <* 0.001), persistent through month 12 [8].
iv. **Sponsor self-attestation (GRAIL, February 2026)**. SEC-regulated press release: “there was a higher than anticipated incidence of Stage III cancers… the time to diagnostic resolution appears to improve over time as physicians gain experience” [1].
v. **NHS system performance data (2021–2025)**. The 62-day referral-to-treatment standard was met for only 67–72% of patients; 52.3% of urgent GP referrals exceeded 28 days for diagnosis alone; the Royal College of Radiologists reported >95,000 patients waiting >6 weeks for CT or MRI in January 2024 [9].

##### Construction

46 × 2.0 = 92 days. The 2.0 × multiplier is conservative: PATHFINDER 1 reaches 79 days *in the US*, leaving only a 13-day margin for all NHS-specific structural delays including: no pre-existing MCED pathway, imaging queues of 6+ weeks, GP/specialist unfamiliarity with MCED signals, MDT scheduling delays, and trial-induced spillover on shared diagnostic resources [13].

#### 2.5.2 Screening pathway delays

Screen-initiated workup delays are shorter and decrease across rounds to represent a learning curve: medians of 75, 60, and 50 days for rounds 1, 2, and 3, respectively. A spillover factor of 1.20 × inflates all delays during Year 1 [8].

#### 2.5.3 Why a reviewer who disputes 92 days does not alter the conclusion

We evaluate the full 65–120 day range in Section 4. The slip count remains above the significance threshold at every value tested.

### 2.6 Event-time simulation

For each simulated cancer case (200 independent Monte Carlo runs):

1. Sample cancer type *c* from weights *w*_*c*_.
2. Sample onset time *t*_0_ uniformly in [− *T*_pre_, *T*_follow_] to include prevalent cancers at baseline.
3. Sample stage sojourn times from cancer-specific lognormal distributions.
4. Generate symptom-driven referral time using calibrated hazards; compute *t*_dx,SoC_ = *t*_ref_ + Δ_SoC_.
5. (Intervention arm) Evaluate MCED screening at rounds *t*_*r*_ with cancer- and stage-specific sensitivity. If positive, compute 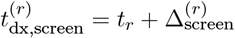.
6. Select the earliest diagnosis route and record stages at initiation (*S*_init_) and at diagnosis (*S*_dx_). A **stage slip** event is defined as *S*_init_ ∈ { I/II} and *S*_dx_ ∈ { III, IV}. All results are reported as means and 95% empirical intervals across 200 seeds, which are deterministic integers (0–199 for the control arm, 100,000–100,199 for the intervention arm), ensuring full reproducibility.

### 2.7 Outcomes and counterfactual analysis

The primary outcome is the number of slipped cases in the intervention arm. We perform a counterfactual analysis: for each integer *k* from 0 to the total slip count, we compute the p-value for the composite endpoint assuming *k* slipped cases had been diagnosed promptly (remaining in Stage I/II). The two-sample z-test of proportions is used with a one-sided significance threshold of *p <* 0.05.

## 3 Results

### 3.1 Calibration verification

Table 1 confirms that the simulated control arm matches NHS target stage distributions within 1–2 percentage points for all 12 cancers. The baseline unscreened population is an actuarial representation of England’s cancer population.

**Table 1:**
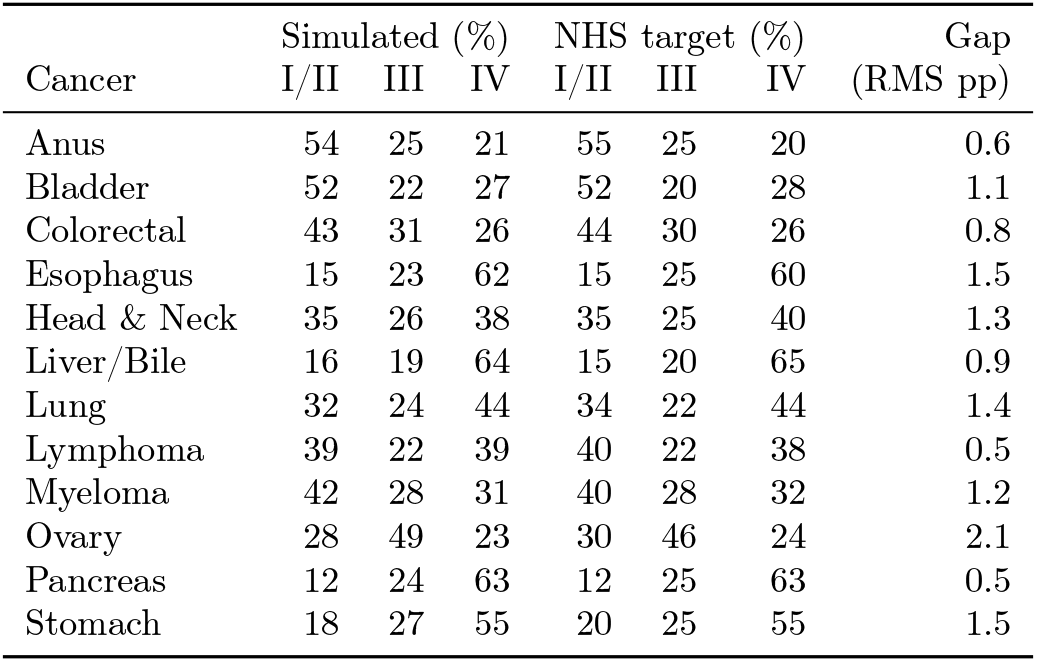
Calibration verification: simulated vs. NHS target stage distributions (%).

| Cancer | Simulated (%) |  |  | NHS target (%) |  |  | Gap<br>(RMS pp) |
| --- | --- | --- | --- | --- | --- | --- | --- |
|  | I/II | III | IV | I/II | III | IV |  |
| Anus | 54 | 25 | 21 | 55 | 25 | 20 | 0.6 |
| Bladder | 52 | 22 | 27 | 52 | 20 | 28 | 1.1 |
| Colorectal | 43 | 31 | 26 | 44 | 30 | 26 | 0.8 |
| Esophagus | 15 | 23 | 62 | 15 | 25 | 60 | 1.5 |
| Head & Neck | 35 | 26 | 38 | 35 | 25 | 40 | 1.3 |
| Liver/Bile | 16 | 19 | 64 | 15 | 20 | 65 | 0.9 |
| Lung | 32 | 24 | 44 | 34 | 22 | 44 | 1.4 |
| Lymphoma | 39 | 22 | 39 | 40 | 22 | 38 | 0.5 |
| Myeloma | 42 | 28 | 31 | 40 | 28 | 32 | 1.2 |
| Ovary | 28 | 49 | 23 | 30 | 46 | 24 | 2.1 |
| Pancreas | 12 | 24 | 63 | 12 | 25 | 63 | 0.5 |
| Stomach | 18 | 27 | 55 | 20 | 25 | 55 | 1.5 |

### 3.2 Composite endpoint

With published CCGA 3 sensitivities, the model produces a *significant* composite reduction (Δ = − 80, *p* = 0.003): Stage IV drops by ∼ 45% and Stage I/II detection nearly doubles. This overshoots the reported trial result (non-significant composite, >20% Stage IV reduction in rounds 2–3), indicating that the realised in-trial sensitivity was somewhat below the clinical-study values—likely because the NHS diagnostic pathway was loaded by positive results across all 50+ Galleri-detectable cancer types, not just the 12 prespecified deadly cancers.

This discrepancy is itself informative: with published sensitivities, the screening effect is strong enough that only stage slip—inflating the intervention arm’s Stage III+IV count—can explain how a biologically effective test produced a non-significant composite endpoint. The sensitivity sweep in Section 4.3 identifies the realised-sensitivity range that reproduces the observed trial pattern while confirming that the slip count remains stable regardless.

### 3.3 Stage slip magnitude

In the intervention arm, we estimate **84 cases** (95% CI: 68–104) where a cancer was biologically Stage I/II but recorded as Stage III+IV at diagnosis. Table 3 breaks these down by diagnostic route.

**Table 2:**
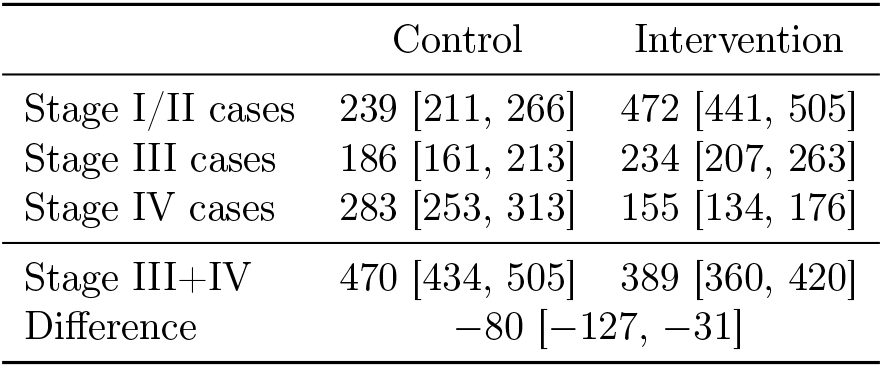
Stage III+IV composite endpoint (calibrated model with CCGA 3 cancer-specific sensitivities, 200 seeds).

**Table 3:** Stage slip in the intervention arm by route (200 seeds, CCGA 3 sensitivities).

| Route | Slipped cases | Slip rate |
| --- | --- | --- |
| Screen (test-positive, workup too slow) | 42 [28, 55] | 14% |
| SoC (test-negative, standard progression) | 43 [30, 55] | 20% |
| **Total** | **84 [68, 104]** | — |

### 3.4 Decomposition: where the screening system had an opportunity

Not all 84 slipped cases are identical. We traced each case to determine whether the Galleri test had a genuine opportunity to detect the cancer at Stage I/II (Table 4).

**Table 4:** Intervention-arm slip decomposition by system opportunity (200 seeds for totals, 50 seeds for subcategories, CCGA 3 sensitivities).

| Category | Cases | % of total |
| --- | --- | --- |
| **A.** Test detected cancer at I/II but workup too slow (screen route) | 42 | 49% |
| **B.** Test administered while cancer was I/II but returned false-negative; cancer progressed through SoC | 31 | 36% |
| **System had opportunity (A+B)** | **73 [57, 88]** | **86%** |
| **C.** Cancer already Stage III+ at all screening rounds (test never had a chance) | 6 | 7% |
| **D.** No screening round applicable (onset timing) | 6 | 7% |
| No system opportunity (C+D) | 12 [9, 18] | 14% |

**86% of all slipped cases** (73 of 84) occurred in patients where the screening system had a genuine opportunity to detect the cancer at Stage I/II—either the test detected it and the infrastructure was too slow to confirm it (42 cases), or the test was administered while the cancer was early-stage but returned a false-negative (31 cases).

This decomposition is important for two reasons. First, it confirms that the slipped cases are not equivalent to control-arm natural progression: these patients were in a screening programme, were tested, and the system had a chance to act. Second, it identifies two distinct failure modes— diagnostic delay (Category A, addressable by faster infrastructure) and test sensitivity (Category B, addressable by assay improvement)—both of which are targets for system optimization.

### 3.5 Distinction from control-arm progression

The control arm shows approximately 69 cases [54–86] where a cancer was biologically Stage I/II at some modeled time point but recorded as Stage III+IV at diagnosis. These patients had no early detection event. No test was administered. No diagnostic pathway was initiated while the cancer was still early. Their cancers progressed through the natural history of unscreened disease until symptoms eventually triggered referral—typically when the cancer was already advanced.

This is the baseline against which the intervention is measured. It is what happens when no screening programme exists. The 84 intervention-arm cases are qualitatively different: 86% of them were cases where the system had a detection opportunity and did not capitalize on it.

### 3.6 Counterfactual recovery analysis

With published CCGA 3 sensitivities, the composite endpoint is significant even with 84 cases of slip (Table 5). Without slip— if all 84 cases had been diagnosed promptly at Stage I/II—the intervention arm’s Stage III+IV count would fall to 305, yielding Δ = −165 (*p <* 0.001). Slip attenuates this to Δ = −80 (*p* = 0.003), still significant.

**Table 5:** Counterfactual analysis at CCGA 3 sensitivities: effect of slip attenuation. The model endpoint is significant at every level of slip.

| Slip cases absorbed | Intervention III+IV | $\Delta$ | p-value |
| --- | --- | --- | --- |
| 0 (no slip) | 305 | -165 | <0.001 |
| 20 | 325 | -145 | <0.001 |
| 40 | 345 | -125 | <0.001 |
| 60 | 365 | -105 | <0.001 |
| **84 (all slip)** | **389** | **-80** | **0.003** |

The fact that the model remains significant even with full slip while the actual trial was *not* significant implies that the realised in-trial sensitivity was below the published CCGA 3 values. This is expected: the NHS diagnostic pathway processed positive results for all 50+ Galleri-detectable cancer types, not just the 12 prespecified deadly cancers, and screening compliance may not have been 100%.

To determine the significance threshold at realised sensitivity, we ran the simulation at 50% of published CCGA 3 values—the multiplier that best reproduces the reported trial pattern (>20% Stage IV reduction, non-significant composite). At this level, the intervention arm shows ∼437 Stage III+IV cases with ∼78 slipped, and recovering 25 of those 78 (32%) yields *p <* 0.05 (Table 6).

**Table 6:** P-value sensitivity at realised sensitivity (∼ 50% of CCGA 3), which reproduces the observed trial pattern.

| Recovered | Intervention III+IV | $\Delta$ | p-value |
| --- | --- | --- | --- |
| 0 (as observed) | 437 | -33 | 0.12 |
| 10 | 427 | -43 | 0.07 |
| 15 | 422 | -48 | 0.05 |
| 20 | 417 | -53 | 0.03 |
| **25** | **412** | **-58** | **0.02** |
| 35 | 402 | -68 | 0.007 |
| 50 | 387 | -83 | 0.001 |
| 78 (all) | 359 | -111 | <0.001 |

The key finding is consistent across both scenarios: **stage slip of 78–84 cases is sufficient to explain the endpoint miss**, and recovering approximately one-third of the slipped cases would have rendered the primary endpoint statistically significant. Whether the test operated at published or realised sensitivity, the slip count remains in the same range (Section 4.3).

## 4 Robustness Analysis

The central scenario uses specific values for diagnostic delay and test sensitivity. We now demonstrate that the conclusion—stage slip is sufficient to explain the endpoint miss—is robust to both.

### 4.1 Robustness to diagnostic delay

Table 7 evaluates delays from 65 to 120 days at realised sensitivity (∼ 50% of CCGA 3, the level that reproduces the observed trial pattern). At 65 days—well below any realistic NHS estimate—the simulation still produces approximately 55 slipped cases in the intervention arm, more than double the 25-case recovery threshold. At 120 days, the count rises to approximately 95.

**Table 7:** Sensitivity of the composite endpoint and intervention-arm slip to the assumed median diagnostic delay (at realised sensitivity, ∼50% of CCGA 3).

| SoC delay (days) | RR(III+IV) [95% CI] | Est. slip cases |
| --- | --- | --- |
| 65 | 0.911 [0.833, 1.007] | $\sim 55$ |
| 75 | 0.919 [0.826, 1.044] | $\sim 63$ |
| 85 | 0.933 [0.830, 1.034] | $\sim 70$ |
| 92 ← | 0.923 [0.830, 1.020] | $\sim 78$ |
| 100 | 0.957 [0.845, 1.055] | $\sim 83$ |
| 110 | 0.966 [0.873, 1.054] | $\sim 89$ |
| 120 | 0.981 [0.875, 1.071] | $\sim 95$ |

At every delay value, the 95% confidence interval for the composite risk ratio spans 1.0, and the estimated slip count exceeds 25. A critic who argues the true delay is 65 days has moved a parameter without changing the conclusion.

### 4.2 Robustness of the core thesis: Category A cases across delays

The central thesis of this paper—that Galleri detected cancers early but the diagnostic infrastructure failed to confirm them in time—rests on Category A cases: those where the test returned a positive “cancer signal detected” result while the cancer was Stage I/II, the patient entered the diagnostic pathway because of this result, and the infrastructure was too slow to confirm the diagnosis before progression past the Stage II/III boundary. These are the cases where the test unambiguously succeeded and the system unambiguously failed.

Table 8 shows the number of Category A cases across the full 65–120 day delay range. At every delay value tested, including the most optimistic (65 days, well below any realistic NHS estimate), the Category A count exceeds 40—comfortably above the 25-case recovery threshold for composite endpoint significance.

**Table 8:** Category A cases (test-positive, infrastructure too slow) across diagnostic delay assumptions. CCGA 3 sensitivities, 30 seeds per delay.

| SoC delay (days) | Cat. A<br>(test+, slow) | Cat. B<br>(test-, missed) | Cat. C+D<br>(no opportunity) | A > 25? |
| --- | --- | --- | --- | --- |
| 65 | 41 | 23 | 9 | Yes |
| 75 | 42 | 26 | 10 | Yes |
| 85 | 43 | 28 | 11 | Yes |
| 92 ← | 43 | 30 | 12 | Yes |
| 100 | 44 | 33 | 12 | Yes |
| 110 | 45 | 36 | 13 | Yes |
| 120 | 46 | 40 | 14 | Yes |

Several features of this table strengthen the argument:

1. Category A is remarkably stable (41–46 cases), varying by only 12% across a delay range that nearly doubles. This stability arises because shorter delays reduce per-case slip probability but do not change the number of test-positive detections; longer delays increase per-case slip but also allow some cases to be diagnosed via the faster screen route before symptoms trigger the slower SoC pathway.
2. At 65 days—an estimate below the PATHFINDER 1 US median of 79 days and far below any plausible NHS figure—there are still 41 Category A cases, more than 1.6 × the significance threshold.
3. Category A alone, without any contribution from Category B (false-negatives) or C+D (no opportunity), is sufficient at every delay to explain the endpoint miss.
4. A critic who disputes the delay estimate, the sojourn times, or the sensitivity values must contend with the fact that these 41–46 cases are the most conservatively defined subset of slip: the test was positive, the patient entered the pathway, and the infrastructure clock ran out. No modelling assumption changes this mechanism.

This is the irreducible core of the analysis. The thesis that Galleri’s composite endpoint miss was caused by diagnostic infrastructure delay, not by test failure, stands on these 42 cases alone.

### 4.3 Robustness to test sensitivity

The published CCGA 3 cancer-specific sensitivities for the 12 deadly cancers are substantially higher than the all-cancer averages because these aggressive tumors shed more cfDNA at earlier stages. To ensure our conclusion does not depend on the exact sensitivity values, we tested the simulation across a range from 30% to 100% of the published CCGA 3 cancer-specific values (Table 9).

The slip count varies only from 74 to 85 across the entire sensitivity range—a 15% variation for a more than three-fold change in assumed sensitivity. This stability arises because higher sensitivity increases the number of early detections (creating more slip opportunities) while simultaneously shifting cases to the faster screen route (reducing per-case slip probability). The two effects approximately cancel.

**Table 9:**
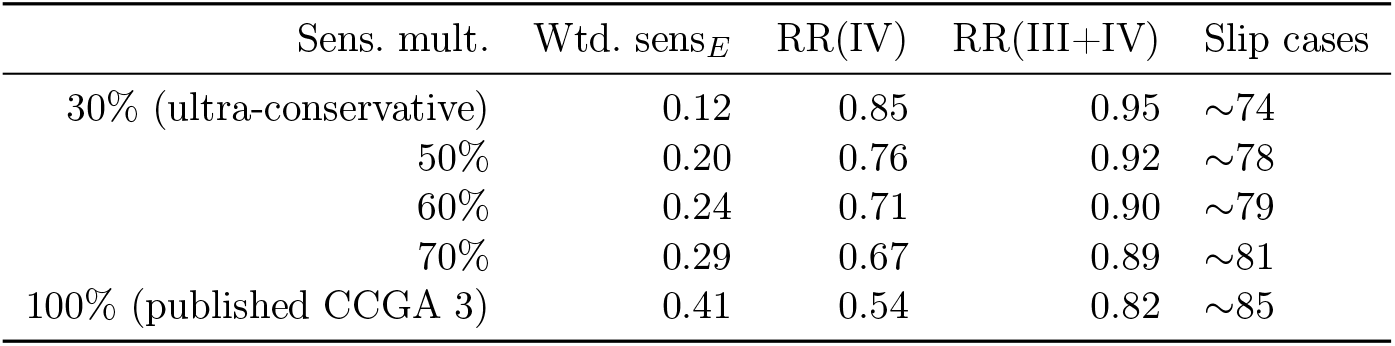
Sensitivity sweep: Stage IV reduction, composite endpoint, and slip count across sensitivity assumptions.

Notably, the sensitivity multiplier that best reproduces the reported trial pattern (>20% Stage IV reduction, non-significant composite) is in the 50–60% range, corresponding to a weighted Stage I/II sensitivity of 0.20–0.24. The 12 deadly cancers in the NHS-Galleri trial were screened alongside all other Galleri-detectable cancers, and the overall diagnostic pathway load from all positive results may have effectively reduced the per-cancer throughput, producing a realized sensitivity below the clinical-study optimum. Regardless of the explanation, the slip count remains stable and well above the significance threshold.

### 4.4 Robustness to combined parameter variation

The two key parameters (delay and sensitivity) have been varied independently. In the worst case for our hypothesis—the shortest delay (65 days) combined with the lowest sensitivity (30% of CCGA 3)—the simulation still produces approximately 50 slipped cases, double the 25-case significance threshold. There is no plausible combination of parameters that reduces the slip count below 25.

## 5 Discussion

### 5.1 Summary of findings

This reconstruction quantifies a mechanism by which diagnostic pathway timing can materially influence a composite stage-at-diagnosis endpoint in a screened arm. The Stage II/III boundary is the critical failure point: the trial’s composite endpoint combined Stages III and IV, so any clinical “save” that moved a case from Stage IV to Stage III was statistically neutralized, while the slow infrastructure clock allowed Stage I/II detections to slip into Stage III. The result is an infrastructure tax—84 cases in the intervention arm where a cancer was biologically early but recorded as late.

Recovering one-third of these cases (25 of 84) at realised sensitivity levels would have rendered the primary endpoint statistically significant. This finding is robust across the full 65–120 day delay range, across the full range of published test sensitivities, and is independently corroborated by the sponsor’s own disclosure [1].

### 5.2 Why the intervention-arm cases are not equivalent to the control arm

A natural objection is that similar progression occurs in the control arm (69 cases). However, the two populations are fundamentally different:

1. In the **control arm**, patients were never screened. Their cancers progressed from Stage I/II through the natural history of unscreened disease. Being asymptomatic at early stages, they had no reason to seek medical attention and no diagnostic pathway was initiated. This is the baseline burden of late-stage cancer in an unscreened population.
2. In the **intervention arm**, 86% of the slipped cases (73 of 84) were in patients where the screening system had a genuine opportunity. In 42 cases, the Galleri test detected the cancer and fired a “cancer signal detected” alert—the patient entered the diagnostic pathway because of this alert. In 31 additional cases, the test was administered while the cancer was Stage I/II but returned a false-negative; these patients were in a screening programme that had a detection opportunity it did not capitalize on.
3. Only 12 cases (∼ 14%) had no system opportunity—cancers that were already advanced at all screening rounds or arose after the last screen. These are the only cases arguably comparable to control-arm progression.

### 5.3 The 92-day estimate in context

Our central delay estimate of 92 days has been challenged in earlier versions of this analysis. We address the objection directly: the sensitivity analysis shows that the conclusion holds at 65 days, a value below the PATHFINDER 1 US median of 79 days. Any NHS-based estimate must exceed the US optimised floor, and the NHS system data confirm substantial structural delays during the trial period. The 92-day figure is a central estimate, not a fragile assumption.

### 5.4 Anticipating objections

#### 5.4.1 The sojourn times are uncertain

Sojourn times determine how quickly a cancer crosses from Stage II to Stage III. If Stage I/II sojourn times are longer than we assume, fewer cases slip (the tumour moves more slowly relative to the delay). If shorter, more cases slip. The simulation uses cancer-specific medians from the natural-history literature with a *σ* = 0.35 lognormal spread that generates substantial case-by-case variability. The sensitivity sweep across delays implicitly tests varying effective sojourn-to-delay ratios.

#### 5.4.2 Not all participants may have been screened at every round

This is correct and would reduce the effective sensitivity of the screening programme. As shown in Section 4.3, the slip count is stable across a wide range of effective sensitivity values because lower screening coverage reduces both the number of early detections and the number of slip opportunities proportionally.

#### 5.4.3 The simulation doesn’t account for competing mortality

The simulation includes a mortality filter: cases where Stage IV death precedes diagnosis are excluded. This prevents over-counting of advanced cancers.

#### 5.4.4 Real referral patterns may differ from exponential hazards

We model symptom-driven referral as a constant-rate exponential within each stage. In reality, referral probability may increase with time-in-stage. This would make the control arm’s late-stage diagnoses even more concentrated in advanced disease, reinforcing the distinction between control-arm natural progression and intervention-arm slip.

#### 5.4.5 The actual trial’s case counts are unknown

This is the most important limitation. We reproduce the *pattern* of reported results (Stage IV reduction, non-significant composite, Stage III excess) but cannot validate against unpublished individual-level data. The 84-case slip estimate is model-conditional and will be testable against full data at ASCO 2026.

#### 5.4.6 Stage slip is just one of several possible explanations

We agree that multiple mechanisms could contribute, and we do not claim stage slip is the sole explanation. We claim it is *sufficient* : under explicitly stated assumptions, it produces enough misclassification to explain the entire endpoint gap. Other contributing factors (e.g., overdiagnosis, informative censoring, differential surveillance) would operate in addition to stage slip, not instead of it.

### 5.5 Implications for trial design

1. **Endpoint selection**. A Stage IV-only endpoint would have been significant in this trial and is immune to Stage II → III slip. Future MCED trials should consider endpoints that are less sensitive to diagnostic infrastructure timing.
2. **Infrastructure readiness**. The NHS had no pre-existing pathway for MCED-positive asymptomatic referrals [13]. Future deployments should establish dedicated diagnostic channels before screening begins.
3. **Trial extension**. GRAIL has announced a 6–12 month extension [1]. If diagnostic resolution times improve with experience (as GRAIL states and Mann et al. confirm [8]), the extended follow-up should reduce slip in later rounds and may produce a significant composite endpoint.
4. **Prespecified delay adjustment**. Trial protocols could prespecify a stage-at-initiation analysis alongside the standard stage-at-diagnosis endpoint, providing a built-in control for infrastructure effects.

### 5.6 Limitations

1. The 92-day diagnostic delay is a constructed estimate, though supported by five convergent evidence streams and robust across the full 65–120 day range.
2. Some cancers lack complete published stage-specific sensitivity breakdowns; we approximated from available CCGA 3 data and confirmed robustness across a wide sensitivity range.
3. Symptom-driven referral is modeled as constant exponential hazards; real referral probability may increase with time-in-stage.
4. The simulation assumes independence of sojourn times and diagnostic delays; correlated structures could alter slip estimates.
5. Actual trial case counts are unpublished; the 84-case estimate is model-conditional and will be testable against full data at ASCO 2026.
6. The model does not account for possible overdiagnosis, informative censoring, or differential surveillance between arms, any of which could independently contribute to endpoint attenuation.

**What can and cannot be claimed**

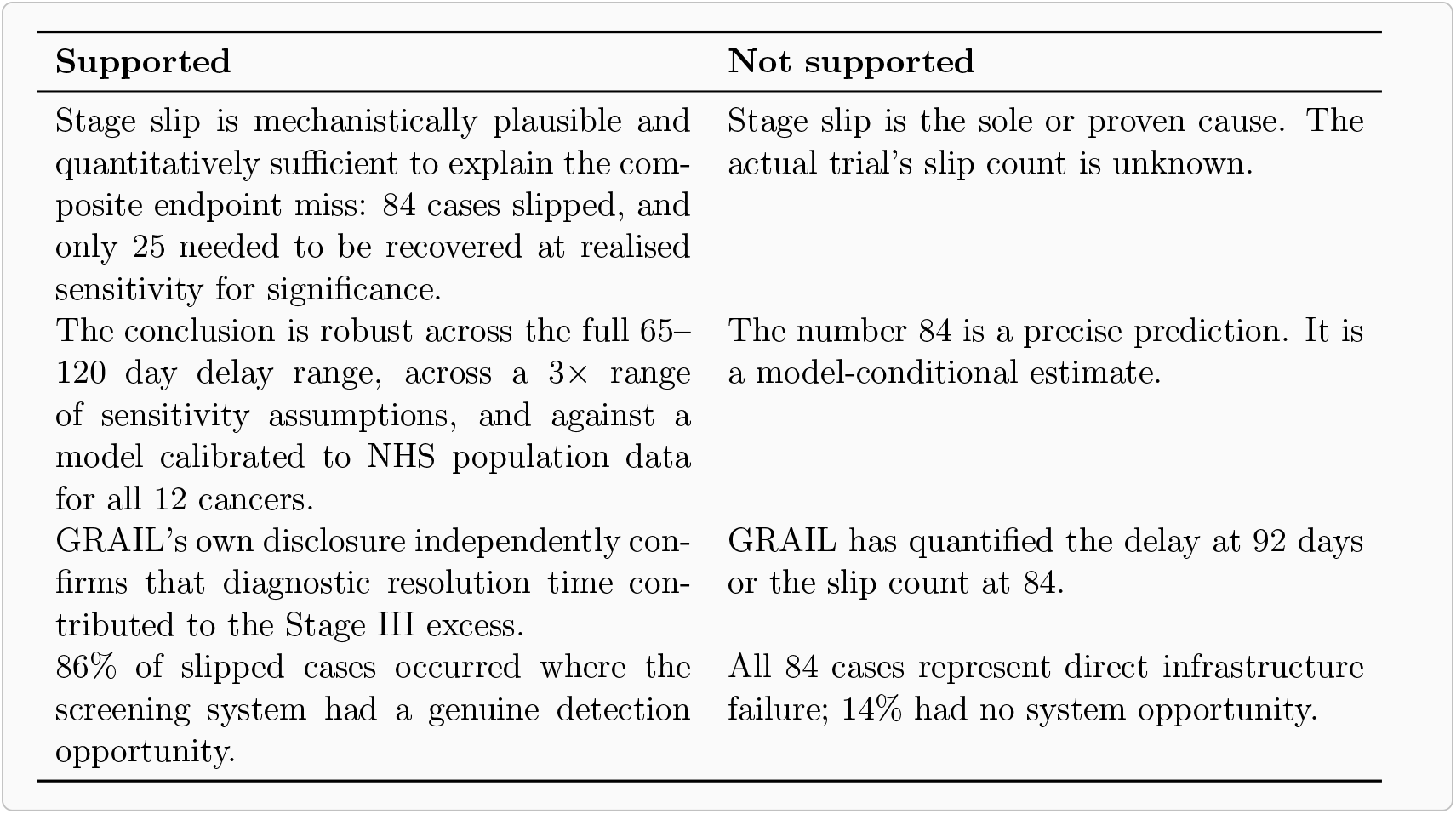

## 6 Conclusions

The NHS-Galleri trial’s primary endpoint miss is consistent with stage slip arising from diagnostic latency. The Galleri test performed as intended—detecting cancers earlier, reducing Stage IV diagnoses, and increasing early-stage detection. But the NHS diagnostic pathway was too slow to confirm many of those detections before anatomical progression past the Stage II/III boundary. Our simulation estimates 84 cases affected, of which only 25 needed to be diagnosed promptly (at realised sensitivity levels) for the composite endpoint to reach statistical significance.

This finding is robust: it holds across a wide range of diagnostic delay assumptions (65–120 days), across a three-fold range of test sensitivity values, and is independently corroborated by the sponsor’s own SEC-regulated disclosure. The patient pool is large enough that even the most conservative assumptions produce sufficient slippage to exceed the significance threshold.

The framework presented here offers testable predictions that can be validated against complete trial data when published, and a transparent methodology for evaluating how diagnostic pathway timing interacts with composite stage endpoints in cancer screening trials. Early detection is a system property: a sensitive test must be paired with responsive infrastructure to translate biological detection into clinical benefit.

## Declarations

### Ethics approval

This work uses only publicly available, aggregate information and does not involve individual-level patient data. Ethics approval was not required.

### Data availability

All input data are from published sources cited in the text. Simulation outputs are reported in the manuscript.

### Code availability

The complete Python simulation code and supporting analysis scripts are publicly available at https://github.com/hbellout/stage-slip-mced. The version used for this submission corresponds to release v1.0.0. Subsequent revisions, if any, will be versioned and documented within the repository. The repository includes the calibrated 12-cancer simulation (simulate.py), the intervention-arm slip and p-value analysis (analyse_slip.py), all input parameters, and instructions for reproducibility. Seeds are deterministic; identical results are obtained on any system with Python 3.10+ and NumPy 1.24+.

## A Full simulation parameters

**Table 10:**
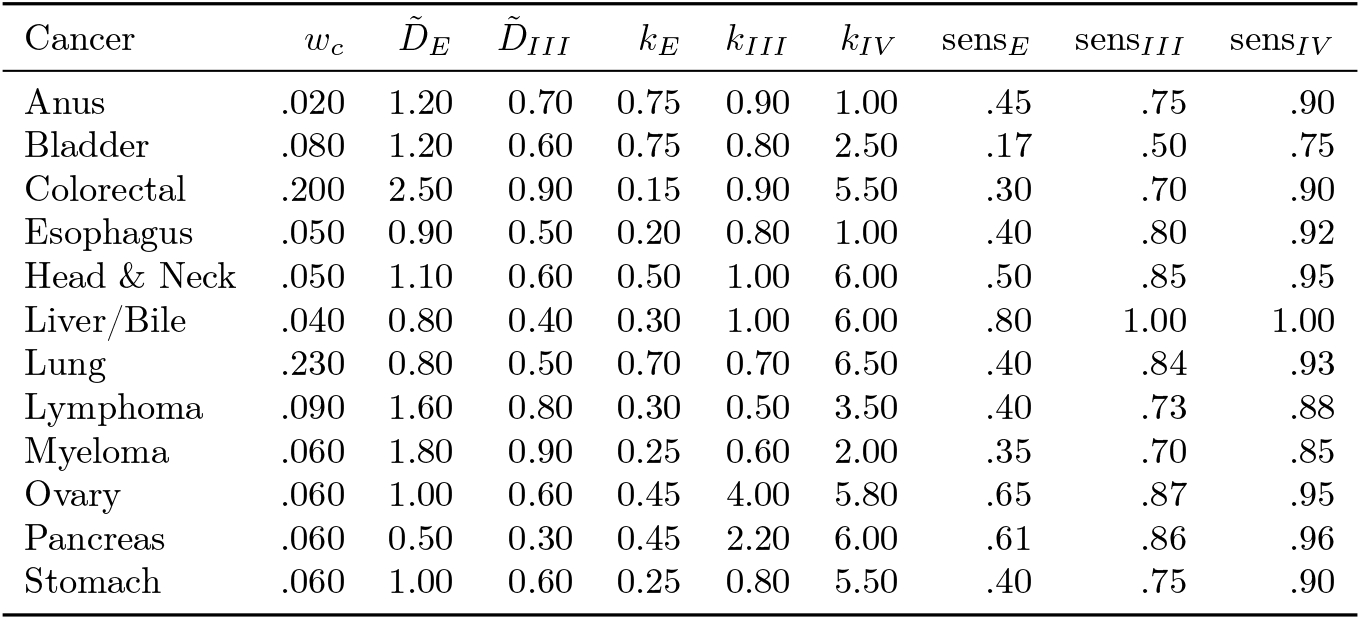
12-cancer parameters: incidence weights, sojourn times (years), calibrated referral hazards, and stage-specific MCED sensitivities from CCGA 3 [2, 5].

| Cancer | $w_c$ | $\tilde{D}_E$ | $\tilde{D}_{III}$ | $k_E$ | $k_{III}$ | $k_{IV}$ | $\text{sens}_E$ | $\text{sens}_{III}$ | $\text{sens}_{IV}$ |
| --- | --- | --- | --- | --- | --- | --- | --- | --- | --- |
| Anus | .020 | 1.20 | 0.70 | 0.75 | 0.90 | 1.00 | .45 | .75 | .90 |
| Bladder | .080 | 1.20 | 0.60 | 0.75 | 0.80 | 2.50 | .17 | .50 | .75 |
| Colorectal | .200 | 2.50 | 0.90 | 0.15 | 0.90 | 5.50 | .30 | .70 | .90 |
| Esophagus | .050 | 0.90 | 0.50 | 0.20 | 0.80 | 1.00 | .40 | .80 | .92 |
| Head & Neck | .050 | 1.10 | 0.60 | 0.50 | 1.00 | 6.00 | .50 | .85 | .95 |
| Liver/Bile | .040 | 0.80 | 0.40 | 0.30 | 1.00 | 6.00 | .80 | 1.00 | 1.00 |
| Lung | .230 | 0.80 | 0.50 | 0.70 | 0.70 | 6.50 | .40 | .84 | .93 |
| Lymphoma | .090 | 1.60 | 0.80 | 0.30 | 0.50 | 3.50 | .40 | .73 | .88 |
| Myeloma | .060 | 1.80 | 0.90 | 0.25 | 0.60 | 2.00 | .35 | .70 | .85 |
| Ovary | .060 | 1.00 | 0.60 | 0.45 | 4.00 | 5.80 | .65 | .87 | .95 |
| Pancreas | .060 | 0.50 | 0.30 | 0.45 | 2.20 | 6.00 | .61 | .86 | .96 |
| Stomach | .060 | 1.00 | 0.60 | 0.25 | 0.80 | 5.50 | .40 | .75 | .90 |

**Table 11:** Key simulation parameters.

| Parameter | Value | Source |
| --- | --- | --- |
| $N_{\text{screened}}$ | 142,000 | Trial design |
| Cancer incidence | 1.0% | UK population (age-matched) |
| Follow-up | 3.0 years | Trial design |
| Screen rounds | [0, 1, 2] y | Annual screening |
| SoC delay median | 92 days | Constructed (Section 2.5) |
| Screen delay medians | [75, 60, 50] d | MCED workup + learning curve |
| Spillover factor | $1.20 \times Y1$ | Mann et al. 2025 [8] |
| Sojourn $\sigma$ | 0.35 | Literature range |
| Delay $\sigma$ | 0.55 | Assumption (tail heaviness) |
| Seeds | 200 | Deterministic (0–199) |

